# Characterization of pretreatment cachexia through cytokine and nutritional analysis in patients with non-small cell lung carcinoma: the Marató cohort

**DOI:** 10.1101/2025.11.12.25339907

**Authors:** Sara Hijazo-Pechero, Inmaculada Peiró, Lorena Arribas, Aina Llenas-Bladé, Felipe Jiménez, Joaquim Moreno-Caceres, Fedra Luciano-Mateo, Matías Fernández-Huarte, Nadia Gómez-Serra, Ana Regina González-Tampán, Jesús Brenes, Miguel Mosteiro, Marina Domingo, Alicia Madurga, Andrés Cuéllar, Arturo Navarro-Martin, Míriam Núñez Fernández, Ramón Palmero, Samantha Aso, Susana Padrones, María Jové, David Cordero, Eduard Montanya, Cristina Muñoz-Pinedo, Ernest Nadal

## Abstract

Cancer cachexia is a frequent complication of patients diagnosed with non-small cell lung cancer (NSCLC) that affects their quality of life and their prognosis. A detailed, multidimensional characterization of patients with cancer cachexia is lacking. We conducted a prospective study of 52 patients diagnosed with locally advanced unresectable NSCLC who underwent concurrent chemoradiotherapy followed by immunotherapy. At baseline, 53% of patients met the Fearon’s criteria for cachexia. Cachexia was significantly associated with tumor stage (p < 0.001), performance status (p = 0.027), lower skeletal muscle index (p = 9e-4) and total adipose tissue index (p = 0.004), elevated C-Reactive Protein blood levels (p = 0.004), moderate to severe malnutrition (p < 0.001), reduced caloric intake (p = 0.007) and diminished physical strength (p = 0.001). Proteomic profiling using the O-link platform revealed known and novel biomarkers of cachexia, including inflammatory cytokines. Cachexia was significantly associated with elevated serum levels of CCL23, IL-6, CSF1, complement factor C1QA, motilin, agouti-related protein (AGRP) and soluble CD4 and CD276, and with lower RANKL, CKAP4, LRRN1, FLT3LG or Persephin, among others. These proteins showed varying degrees of correlation with caloric intake. GDF-15 was associated with weight loss (p = 0.00014), but not with cachexia (p=0.08), calorie intake or body composition. This prospective study demonstrates that cancer cachexia is highly prevalent in patients with unresectable locally advanced NSCLC and is associated with specific, understudied cytokines. These findings provide further insight into the biological characterization cachexia and may help identify novel targets for therapeutic interventions.

## INTRODUCTION

Patients with cancer often experience tumor-induced metabolic alterations that, along with anorexia, fatigue, anhedonia, involuntary weight loss, skeletal muscle and adipose tissue wasting, contribute to a multifactorial syndrome known as cancer cachexia. This condition is difficult to assess clinically, worsens prognosis and quality of life, and is associated to poorer cancer treatment tolerance.

Lung cancer is the leading cause of cancer-related mortality worldwide (1). The most common subtype, non-small cell lung carcinoma (NSCLC), is frequently associated with cachexia, which affects approximately 30-45% of patients (2). Cachectic patients are less likely to receive standard treatments or participate in clinical trials due to severe functional decline and disability. Although multiple definitions of cachexia exist, the most widely adopted criteria are those proposed by Fearon et al., which defined cancer cachexia as 1) a body mass index (BMI) < 20□kg/m² with > 2% weight loss, or 2) > 5% involuntary body weight loss over the past 6 months, or 3) sarcopenia (muscle loss) with > 2% weight loss (3).

The analysis of circulating biomarkers may unravel novel therapeutic targets that mediate cancer cachexia. Recent studies showed that the cytokine Growth Differentiation Factor 15 (GDF-15) is consistently elevated in patients with cachexia. Monoclonal antibodies targeting GDF-15, like ponsegromab, have been investigated in clinical trials to alleviate this condition in various disease settings (4). Moreover, GDF-15 is elevated at relapse in patients with surgically resected NSCLC and correlates with cachexia in this setting (5). However, the association of GDF-15 with pre-treatment cachexia remains unclear. Given the marked interpatient heterogeneity, there is a need to identify additional, specific biomarkers to guide more targeted interventions for cancer cachexia. Previous studies investigating cytokine mediators of cancer cachexia have largely relied on preclinical models. In the current study, we conducted a prospective, multidisciplinary, longitudinal assessment of the cachexia phenotype in a cohort of patients with unresectable, locally advanced NSCLC. We evaluated the prevalence of cachexia in this clinical setting at baseline and identified novel candidate mediators of cachexia through proteomic profiling of circulating cytokines.

## METHODS

### Study design and eligibility

This is a prospective observational study that enrolled 52 patients diagnosed with locally advanced NSCLC who were considered medically inoperable or unresectable by the Multidisciplinary Tumor Board of the HUB-ICO Comprehensive Cancer Center. All patients were identified at the first contact with the medical oncologist in the Thoracic Tumors Unit between April 2022 and March 2024. The study protocol and the informed consent were approved by the Bellvitge Ethical Committee (PR116/19) and all patients provided written informed consent. Initially 61 patients were recruited, but only 52 met the study inclusion criteria and 9 could not be included in this study cohort for diverse reasons (see Flow diagram, Sup. Fig. 1).

All patients received platinum-doubled chemotherapy concurrently with thoracic radiotherapy with curative intent, followed with consolidation with durvalumab in patients whose tumors had positive PD-L1 expression and did not have any contraindication to receive immunotherapy. Volumetric Modulated Arc Therapy was administered concurrently from the first day of chemotherapy up to a total dose of 60–66□Gy in daily fractions of 2□Gy over 6 weeks. A 4D thoracic CT scan and a contrast-enhanced thoracic CT scan were performed and fused. The 4DCT scan was used to delineate the internal target volume (ITV) of the tumor, while the contrast-enhanced CT scan improved the accuracy of tumor and nodal station contouring. The gross tumor volume (GTV) included the primary lung tumor and nodal GTV was used to include involved lymph node stations. Only lymph nodes with suspicious FDG uptake, pathological findings on CT, or histological confirmation of malignancy were included in the GTV (selective nodal irradiation). Organs at risk, such as the lungs, trachea, spinal cord, and oesophagus were contoured as per international guidelines (6).

Demographic data, smoking history, and relevant comorbidities, including hypertension, dyslipidemia, and diabetes according to the diagnostic criteria established by the American Diabetes Association in 2019 (7) were collected, along with tumor staging, histology including PD-L1 expression, molecular genotype when available, and treatment information. Body composition, blood tests, cytokine quantification and comprehensive nutritional assessments were conducted both before and after chemoradiotherapy (**Figure 1**).

**Figure 1.**
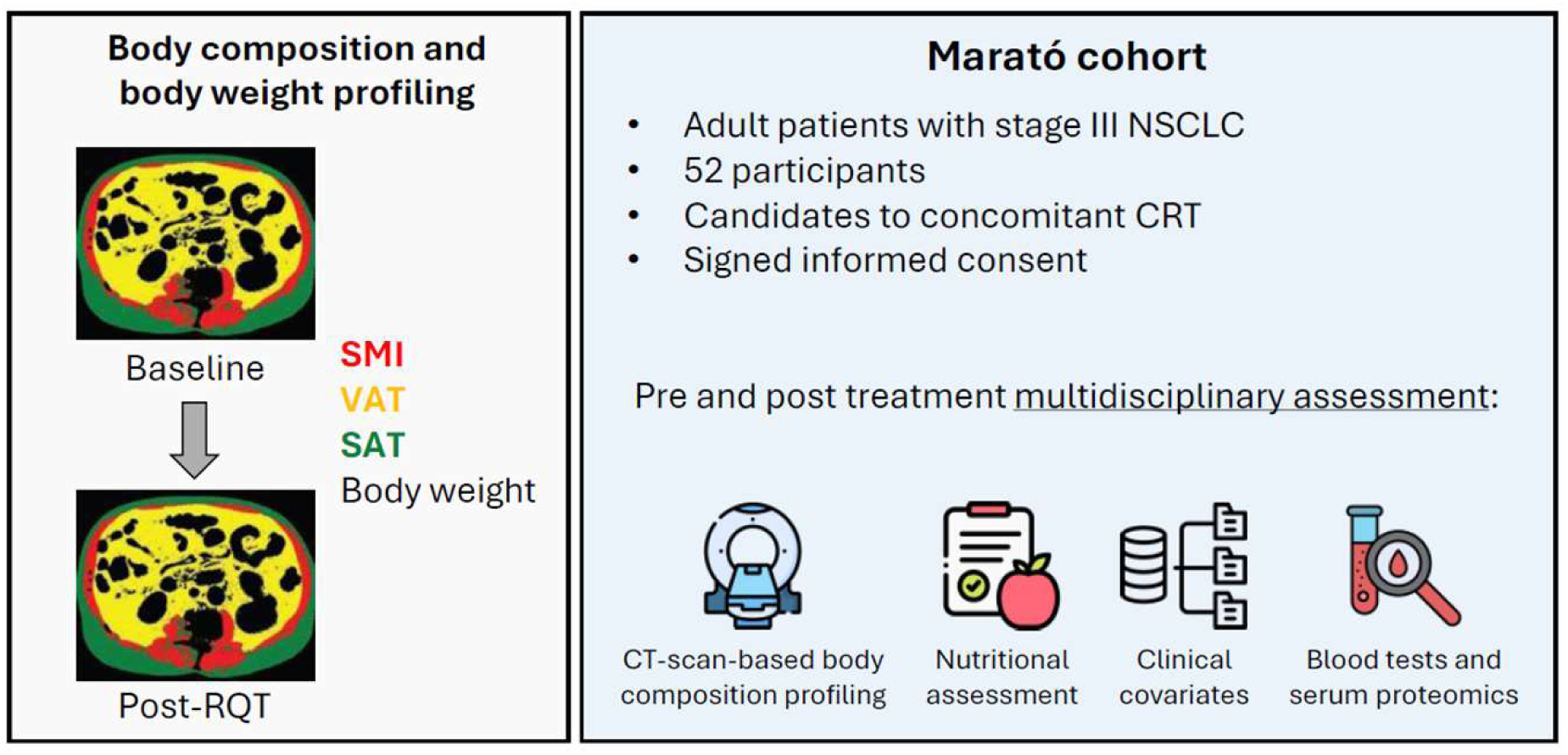
Study overview and body composition profiling workflow. The panel on the left illustrates automated CT-based segmentation of an axial abdominal slice obtained before (Baseline) and after concomitant chemoradiotherapy (Post-CRT). Tissue masks are color-coded—skeletal muscle (SMI, red), visceral adipose tissue (VAT, yellow) and subcutaneous adipose tissue (SAT, green)—and were used, together with body weight, to quantify treatment-related changes in body composition. The right panel summarizes the *Marató* cohort and the multidimensional assessment schedule.

### Metabolic, nutritional and functional assessment

Metabolic, nutritional, and functional assessments were performed before the initiation of oncological treatment (V0) and at the time of tumour evaluation after completing chemoradiotherapy (V1). Nutritional assessments were conducted by a specialist in oncology nutrition. Blood samples were collected from all patients at both V0 and V1. All patients had either a baseline thoraco-abdominal CT scan or a PET-CT within the 8 weeks preceding oncological treatment and a thoraco-abdominal CT scan at the time of tumour assessment, conducted 4 to 6 weeks after V1.

Nutritional data included weight, height, BMI, percentage of weight loss in the six months prior to diagnosis and current weight. Nutritional status was assessed using the Patient-Generated Subjective Global Assessment (PG-SGA) questionnaire (8,9). Energy requirements were estimated using the Mifflin-St Jeor equation (Mifflin et al., 1990) and protein requirements were determined according to ESPEN guidelines (11). Daily protein and calorie intake were estimated using a 24-hour dietary recall and analysed with Dietsource software (Version 4.0). The type of nutritional support required at each visit was also documented. Caloric deficit was defined as the difference between energy requirements and the current daily caloric intake. Functional status was assessed by 4-Meter Gait Speed Test (usual-pace gait speed calculated by timing the traversal of a standardized 4-meter course and expressing the result in meters per second), and by handgrip strength, measured with a Jamar digital dynamometer following a standardised protocol (12). Measurements were performed on the dominant hand, and the highest value obtained from three consecutive attempts was recorded.

### Body composition analysis

All CT images were systematically reviewed for image quality and adequate anatomical coverage at the L3 vertebral level prior to body composition analysis. A total of three baseline CT scans could not be analyzed for body composition due to the absence of the L3 level (two patients) and one due to poor overall image quality. Body composition analysis was quantified from axial CT images at the third lumbar (L3) vertebra using SliceOMatic© software (v5.0 Rev 8, Tomovision, Magog, Canada). Total cross-sectional area (cm^2^) of skeletal muscle (SMA), visceral adipose tissue (VAT), subcutaneous adipose tissue (SAT) and total adipose tissue (TAT) were segmented by predefined Hounsfield unit (HU) thresholds for each tissue: −29 to +150 HU for SMA, −150 to −50 HU for VAT and −190 to −30 HU for SAT (13). SMA, VAT and SAT were normalized to height squared (m^2^) to calculate skeletal muscle index (SMI), visceral adipose tissue index (VATI), subcutaneous adipose tissue index (SATI) and total adipose tissue index (TATI).

Myosteatosis was defined based on mean skeletal muscle radiodensity, expressed in Hounsfield units (HU), calculated within the segmented muscle area. In accordance with the cut-off values proposed by Martin L (14)

### Cachexia

Cachexia was defined as body-weight loss (WL) >5% in the previous 6 months, or WL >2% in the presence of either a BMI <20□kg□m⁻² or sarcopenia, which was defined as low skeletal muscle index (first tertile of the skeletal muscle index variable calculated independently for males and females). The three patients whose CT scan had poor quality or L3 was not available were included because their weight loss was higher than 5% or lower than 2%.

### Blood tests

Blood samples were collected after at least 9 hours fasting and were immediately frozen and stored at −80°C. Blood test included glycemic status markers (Fasting blood glucose (FBG), glycated hemoglobin (HbA1c), insulin, C-peptide), lipid levels (total cholesterol, triglycerides, high-density lipoprotein cholesterol, low-density lipoprotein cholesterol), hormone profile (Insulin growth factor-1 (IGF-1), thyrotropin and thyroxine) and inflammation markers (C-reactive protein (CRP), serum albumin and neutrophil-to-lymphocyte ratio (NLR). The homeostatic model assessment of insulin resistance (HOMA-IR) index was used to evaluate insulin resistance as follows: (Fasting Insulin [UI/L] × Fasting Glucose [mg/dL]) /405. The Modified Glasgow Prognostic Score (mGPS), which serves as a surrogate marker for systemic inflammation, was determined as follows: Patients with C-reactive protein (CRP) □10 mg/L were assigned 0 points; those with CRP >10 mg/L and albumin □35 g/L were assigned 1 point and patients with CRP >10 mg/L and albumin <35 g/L were assigned 2 points (15).

### Proteomic profiling

Peripheral blood was drawn before initiating (V0) and after completing chemoradiotherapy (V1). The blood was processed for serum and plasma extraction within 1□hour and stored at −80°C. Serum from patients was analysed with O-Link technology using the O-Link Explore 384 panel Inflammation I. To measure GDF-15, patient serum was analysed using the Elecsys® GDF-15 assay ref. 08946809190, Roche Diagnostics) and a Cobas8000 e801 system (Roche Diagnostics).

### Statistics

Descriptive statistics were used to summarise baseline demographic and clinical characteristics. Continuous variables are presented as medians with interquartile ranges (IQRs), and categorical variables as frequencies and percentages. Comparisons between groups were performed using the Mann–Whitney U test for continuous variables and the chi-square or Fisher’s exact test for categorical variables. Correlations between clinical and laboratory parameters were assessed using Spearman’s rank correlation coefficient. A two-sided p-value < 0.05 was considered statistically significant, and p-value was adjusted using the false discovery rate (FDR) method for multiple comparisons. Overall Survival (OS) was defined as the time from date of diagnosis to death from any cause or last visit follow-up. Progression-Free Survival (PFS) was defined as the time from treatment initiation to the date of documented disease progression or death from any cause.

The impact of cachexia on OS and PFS was evaluated using univariate and multivariate Cox proportional hazards model, adjusted for disease stage and performance status (PS). All statistical analyses were conducted using R (version 4.4.05.1). Association of cachexia groups with clinicopathological and nutritional covariates was assessed using ‘comparegroups’ package for R (Subirana et al., 2014).

## RESULTS

### Clinical Characteristics of the Study Population

Fifty-two adult patients diagnosed with locally advanced NSCLC and deemed medically inoperable or unresectable by the Multidisciplinary Tumour Board of the HUB-ICO Comprehensive Cancer Centre were included. Baseline patient characteristics are shown in **Table 1**. The median age was 68 years (interquartile range, IQR, 8.5). A total of 42 (80%) patients were male, and the majority (98%) had a history of smoking. The most common histology was squamous cell carcinoma (67%), followed by adenocarcinoma (27%) or not otherwise specified (6%). PD-L1 expression was □1% in 31 patients (60%) and negative in 12 patients (29%). All patients completed the planned chemoradiation treatment and 22 (42%) subsequently initiated consolidative treatment with durvalumab.

**Table 1.** Study population.

| **Characteristics** | **Overall (n = 52)** |
| --- | --- |
| **Age at diagnosis, median, (IQR)** | 68 (8.25) |
| **Sex, No. (%)** |  |
| Male | 42 (81%) |
| Female | 10 (19%) |
| **Smoking history, No. (%)** |  |
| Never | 1 (2%) |
| Former | 34 (65%) |
| Current | 17 (33%) |
| **ECOG PS, No. (%)** |  |
| 0 | 10 (19%) |
| 1 | 31 (60%) |
| 2 | 9 (17%) |
| Unknown | 2 (4%) |
| **Histology, No. (%)** |  |
| Lung adenocarcinoma | 14 (27%) |
| Lung squamous cell carcinoma | 35 (67%) |
| NOS | 3 (6%) |
| **Stage, No. (%)** |  |
| IIB | 2 (4%) |
| IIIA | 27 (52%) |
| IIIB | 14 (27%) |
| IIIC | 8 (15%) |
| Unknown | 1 (2%) |
| **PD-L1, No. (%)** |  |
| PD-L1 < 1% | 15 (29%) |
| PD-L1 1-49% | 14 (27%) |
| PDL1 >= 50% | 17 (33%) |
| Unknown | 6 (12%) |
| **Durvalumab, No. (%)** |  |
| No | 29 (56%) |
| Yes | 22 (42%) |
| Unknown | 1 (2%) |
| **BMI classification, No. (%)** |  |
| Underweight | 4 (8%) |
| Normal weight | 18 (35%) |
| Overweight | 18 (35%) |
| Obesity | 12 (23%) |
| **PG-SGA, No. (%)** |  |
| Good nutritional status | 25 (48%) |
| Moderate malnutrition | 12 (23%) |
| Severe malnutrition | 14 (27%) |
| Unknown | 1 (2%) |
| **Caloric deficit, No. (%)** |  |
| No caloric deficit | 10 (19%) |
| Caloric deficit | 41 (79%) |
| Unknown | 1 (2%) |

Abbreviations: ECOG PS, Eastern Cooperative Oncology Group Performance Status scale; NOS, Not Otherwise Specified; PG-SGA, Patient-generated subjective global assessment.

With a median follow-up of 18 months, 17 patients (33%) experienced tumour progression and 33 patients (65%) remained alive at the time of the analysis. The median progression-free survival (PFS) was 20 months and the median overall survival (OS) was not reached. The median BMI in all patients was 25.64 (IQR, 7.08).

According to the WHO classification, four patients (8%) were underweight, 18 (35%) had normal weight, and 29 (57%) were classified as overweight or obese. Based on the PG-SGA, 24 patients (48%) had an adequate nutritional status, while 26 (52%) exhibited moderate to severe malnutrition at baseline. Additionally, the majority of patients (80%) presented a caloric deficit before initiating chemoradiotherapy.

### Detection of Cachexia Phenotype prior to Oncological Treatment

At baseline, 53% of patients met the Fearon criteria for cachexia (**Figure 2A**). Females were more likely to have cachexia at baseline (80%) compared to males (46%), although these differences were not significant due to limited statistical power and lower representation of women (**Table 2**). Cachexia was strongly associated with tumor stage (p < 0.001), and there was a trend towards association with ECOG performance status (p = 0.065). The majority of non-cachectic patients (79%) had stage IIIA, while two thirds of cachectic patients (67%) were diagnosed with stage IIIB-C tumors. Primary tumor maximum standardized uptake value (SUV_max_) measured by ^18^F-FDG PET/CT was similar in both groups.

**Figure 2.**
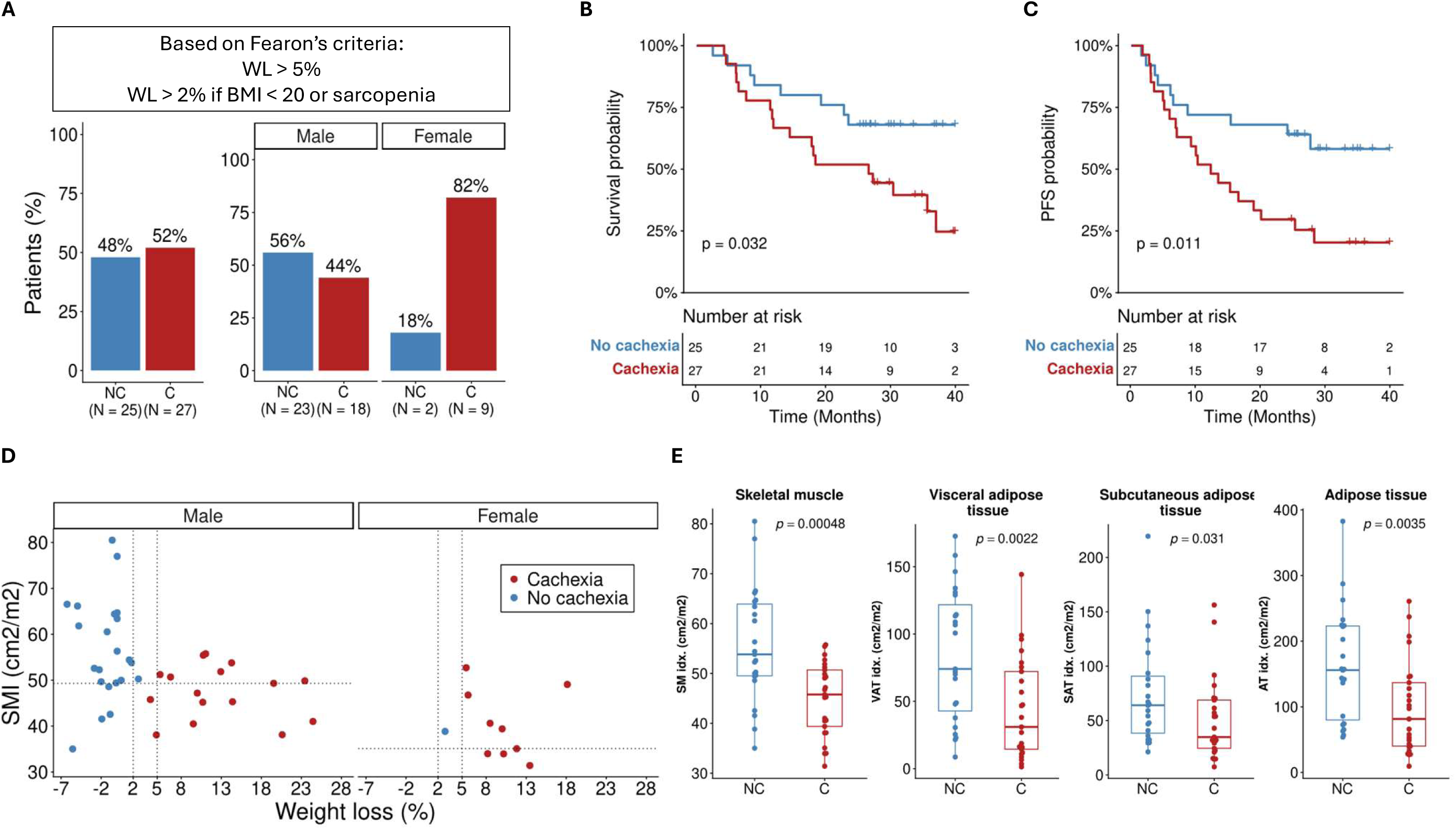
Characterization of the cachexia phenotype in stage III unresectable NSCLC. (A) Prevalence of cachexia according to Fearon’s consensus definition. NC= No Cachexia; C= Cachexia. (B) Kaplan–Meier curve for overall survival (OS). Patients without cachexia (blue) displayed longer OS than those with cachexia (red). (C) Kaplan–Meier curve for progression free survival (PFS). Patients without cachexia (blue) displayed longer PFS than those with cachexia (red). (D) Joint distribution of skeletal-muscle index (SMI, cm²□m⁻²) and percentage of weight loss. Each dot represents an individual patient; colours indicate cachexia status (blue□=□no cachexia, red□=□cachexia), and panels are split by sex. Vertical dashed lines mark Fearon’s weight loss thresholds (5□% or 2□% when BMI□<□20□kg□m⁻² or first SMI index tertile). Horizontal dashed lines denote sex-specific SMI index cut-offs (first tertile). (E) Baseline body-composition differences between cachectic and non-cachectic patients. Box-and-whisker plots show cross-sectional indices for skeletal muscle index (SMI), visceral adipose tissue index (VATi), subcutaneous adipose tissue index (SATI) and total adipose tissue index (TATI). Central line□=□median, box□=□IQR, whiskers□=□1.5×□IQR. p values from two-sided Mann–Whitney U tests are indicated above each comparison.

**Table 2.**
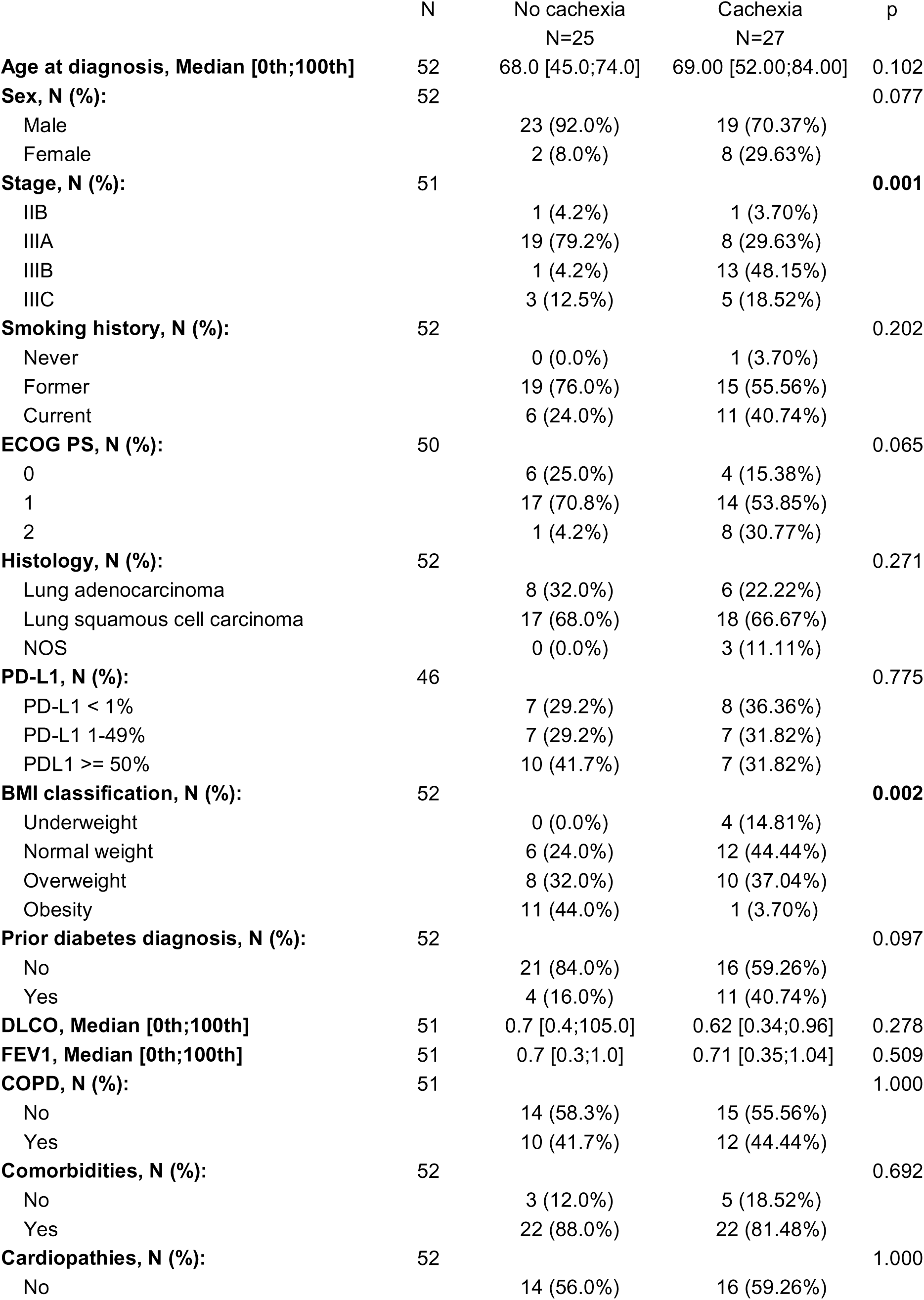
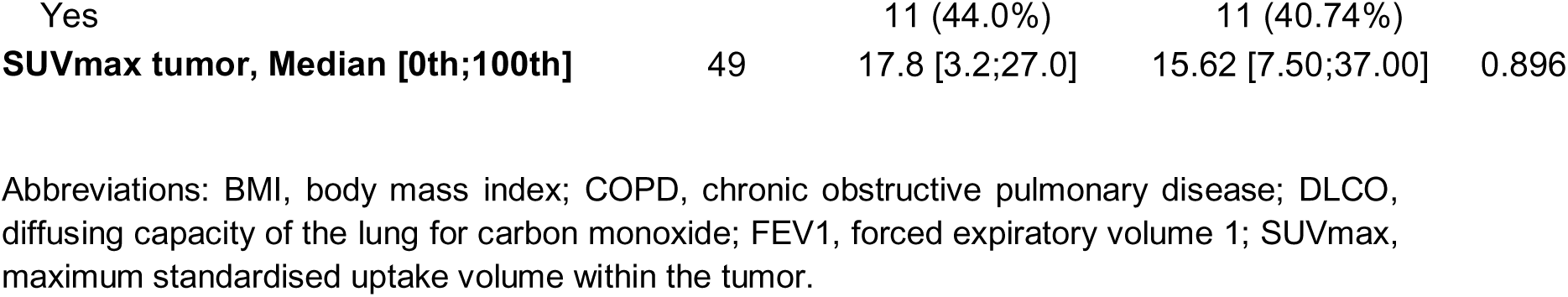
Association of cachexia with clinical features.

We assessed whether cachexia was associated with overall survival (OS) (**Figure 2B**). In the univariate analysis, cachectic patients had a significantly shorter OS compared to non-cachectic patients (univariate HR 2.427; 95% CI, 1.053 – 5.59; p = 0.0373). After adjusting for disease stage and PS, this association was attenuated (adjusted HR 1.63; 95% CI 0.5459 – 4.866; p = 0.3815), suggesting an interaction between this variable and cancer cachexia.

Similarly, in the analysis of progression-free survival (PFS), cachectic patients exhibited a significantly shorter PFS compared to non-cachectic patients (univariate HR 2.588; 95% CI, 1.211 – 5.534; p = 0.0142). However, after adjusting for disease stage and PS, this association was attenuated (adjusted HR 1.9077; 95% CI, 0.7152 – 5.0889; p = 0.197).

The association between cachexia and skeletal muscle mass was further assessed by stratifying patients according to skeletal muscle index (SMI) (**Figure 2D**). Patients were subclassified as low SMI index when the values fell below the first tercile within their sex-specific group. This stratification revealed two phenotypes: cachexia with preserved muscle mass (non-sarcopenic) characterized by normal SMI (top, right, **Figure 2D**), and sarcopenic cachexia, defined by SMI values in the lowest tercile (bottom right, **Figure 2D**). Compared to non-cachectic patients, those with cachexia exhibited significantly lower median SMI (45.8 vs 53.8 cm^2^/m^2^; p=4.8e-4), VATI; 31 vs 74.5 cm^2^/m^2^; p = 0.0022) and SATI; 34.9 vs 64 cm^2^/m^2^; p = 0.031 (**Figure 2E**). The median total adipose tissue index (TATI) was also significantly reduced (81.5 vs 155.9 cm^2^/m^2^, p = 0.0035) in patients with cachexia compared with their counterparts.

### Association Between Cancer Cachexia and Metabolic, Nutritional, and Physical Function Parameters

We evaluated metabolic parameters, total caloric and macronutrient intake (fat, carbohydrate, fiber and protein), nutritional status and physical function to characterize the cachexia phenotype and inform potential nutritional interventions in the future. Cachectic patients had a significantly higher mGSP (p = 0.002) primarily influenced by elevated CRP, and lower fasting insulin levels (p = 0.035) compared with non-cachectic patients (**Table 3**). Cachectic patients consumed significantly fewer calories than non-cachectic individuals (1575.3 kcal/day vs. 1936.1 kcal/day, p = 0.007). Notably, approximately 80% of patients from both groups had a caloric deficit. Regarding macronutrient intake, fat intake was significantly lower in cachectic patients (p = 0.006), while protein intake was comparable between the groups. At baseline, moderate to severe malnutrition was present in 93% of cachectic patients compared to only 5% of patients in the non-cachexia group.

**Table 3.** Metabolic covariates and cachexia.

|  | N | No cachexia<br>N=25 | Cachexia<br>N=27 | p |
| --- | --- | --- | --- | --- |
| Glucose (mg/dl), Median [0th;100th] | 52 | 103.0 [83.0;194.0] | 103.0 [66.0;163.0] | 0.503 |
| Insulin, Median [0th;100th] | 51 | 11.3 [5.0;33.2] | 6.5 [1.7;24.0] | **0.015** |
| HOMA-IR, N (%): | 51 |  |  | 0.330 |
| No insulin resistance (HOMA-IR < 2.5) |  | 11 (44.0%) | 16 (61.5%) |  |
| Insulin resistance (HOMA-IR > 2.5) |  | 14 (56.0%) | 10 (38.5%) |  |
| HbA1c (%), N (%): | 50 |  |  | 0.244 |
| Good glycemic control (< 7%) |  | 22 (88.0%) | 25 (100.0%) |  |
| Fair glycemic control (7-8.5%) |  | 2 (8.0%) | 0 (0.0%) |  |
| Bad glycemic control (> 8.5%) |  | 1 (4.0%) | 0 (0.0%) |  |
| C-Reactive protein (mg/L), N (%): | 51 |  |  | **0.003** |
| RCP < 10 (mg/L) |  | 15 (60.0%) | 4 (15.4%) |  |
| RCP >= 10 (mg/L) |  | 10 (40.0%) | 22 (84.6%) |  |
| Albumin (g/L), N (%): | 52 |  |  | 0.483 |
| Albumin < 35 (g/L) |  | 0 (0.0%) | 2 (7.4%) |  |
| Albumin >= 35 (g/L) |  | 25 (100.0%) | 25 (92.6%) |  |
| mGPS, N (%): | 51 |  |  | **0.002** |
| 0 (RCP <10 (mg/L) plus Alb > 35 (g/L)) |  | 15 (60.0%) | 4 (15.4%) |  |
| 1 (RCP >10 (mg/L) or Alb < 35 (g/L)) |  | 10 (40.0%) | 20 (76.9%) |  |
| 2 (RCP >10 (mg/L) plus Alb < 35 (g/L)) |  | 0 (0.0%) | 2 (7.7%) |  |
| Neutrophil/Lymphocyte ratio, N (%): | 51 |  |  | 0.196 |
| NLR < 3.5 |  | 17 (68.0%) | 12 (46.2%) |  |
| NLR >= 3.5 |  | 8 (32.0%) | 14 (53.8%) |  |
| Caloric intake, Median [0th;100th] | 51 | 1963.4 [1183.3;2902.9] | 1575.3 [643.6;2460.8] | **0.005** |
| Carbohydrates intake, Median [0th;100th] | 51 | 190.2 [101.1;381.3] | 162.0 [88.3;301.9] | 0.126 |
| Protein intake, Median [0th;100th] | 51 | 70.4 [32.4;117.5] | 67.2 [15.8;104.0] | 0.637 |
| Fat intake, Median [0th;100th] | 51 | 89.3 [46.2;194.9] | 66.0 [14.3;106.4] | **0.004** |
| Fiber intake, Median [0th;100th] | 51 | 13.2 [3.3;26.5] | 11.0 [2.0;23.6] | 0.126 |
| PG-SGA, N (%): | 51 |  |  | **<0.001** |
| Good nutritional status |  | 23 (95.8%) | 2 (7.4%) |  |
| Moderate malnutrition |  | 1 (4.2%) | 11 (40.7%) |  |
| Severe malnutrition |  | 0 (0.0%) | 14 (51.9%) |  |
| Caloric deficit, N (%): | 51 |  |  | 1.000 |
| No caloric deficit |  | 19 (79.2%) | 22 (81.5%) |  |
| Caloric deficit |  | 5 (20.8%) | 5 (18.5%) |  |
| Dinamometry, Median [0th;100th] | 52 | 34.1 [19.9;49.8] | 24.8 [14.4;44.2] | **<0.001** |
| Speed test (time), Median [0th;100th] | 50 | 3.7 [2.7;5.9] | 4.2 [2.3;8.3] | 0.113 |
| Speed test (score), Median [0th;100th] | 50 | 4.0 [3.0;4.0] | 4.0 [2.0;4.0] | 0.106 |
| Mioesteatosis, N (%): | 48 |  |  | 0.410 |
| No |  | 11 (47.8%) | 8 (32.0%) |  |
| Yes |  | 12 (52.2%) | 17 (68.0%) |  |

We also evaluated the impact of cachexia on physical performance. While no significant differences were observed in gait-speed tests, cachectic patients exhibited markedly reduced handgrip strength (p = 0.001), consistent with their lower muscle mass. Importantly, these findings remained when analysis were restricted to male participants, acknowledging that many of these parameters can be sex dependent (Supporting Information **Table S1**).

### Pre-treatment GDF-15 correlates with Weight Loss but not with other Cachexia-Related Variables

The median level of pre-treatment GDF-15 was 2182 ng/ml, being above the 1.5 ng/mL threshold used in the clinical trial by Groarke *et al* (4) in 74% of patients. Notably, GDF-15 levels did not differ significantly between patients with cachexia and their counterparts (**Figure 3A**). This suggests that GDF-15 achieved an 80% sensitivity for detecting cachexia but had a low specificity leading to a false-positive rate of 70%.

**Figure 3.**
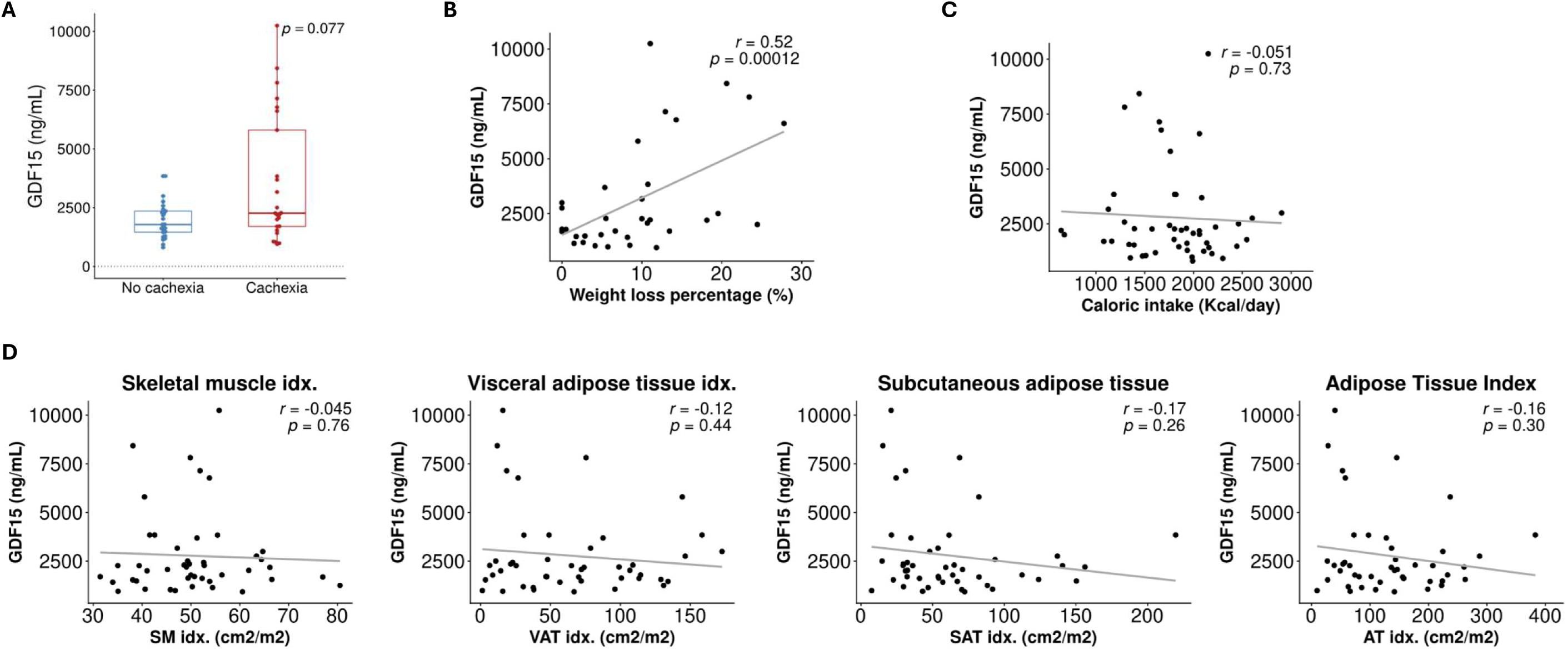
Association of GDF15 with lung cancer cachexia. (A) Plasma GDFl7J15 concentrations (ng ml⁻¹) in patients with and without cachexia; box shows median and IQR, whiskers = 1.5 × IQR (Mann–Whitney U test, p = 0.08). (B) Scatterl7Jplot of GDFl7J15 blood levels versus percentage bodyl7Jweight loss over the preceding 6 months (Spearman r = 0.52, p = 0.00014). (C) Scatterl7Jplot of GDFl7J15 blood levels versus daily caloric intake (kcal day⁻¹) (r = –0.048, p = 0.74). (D) Correlations between GDFl7J15 blood levels and baseline bodyl7Jcomposition indices: skeletall7Jmuscle index (SMI), visceral adipose tissue index (VATI), subcutaneous adipose tissue index (SATI) and total adipose tissue index (TATI); Pearson r and p values shown in each panel. Solid lines in panels B–D represent simple linearl7Jregression fits.

To further explore the relationship between GDF-15 and cachexia-related parameters, we analysed its correlation with weight loss, caloric intake and body composition. Interestingly, we found that GDF-15 levels were significantly correlated with percentage of weight loss (p = 0.00014, r = 0.52; **Figure 3B**), but not with caloric intake (p = 0.74, **Figure 3C**), weight-normalized caloric intake (p=0.70, not shown), or body composition measurements (**Figure 3D**).

### Cytokines Differentially Elevated in Patients with Cachexia

To identify novel circulating biomarkers beyond GDF-15 and uncover potential new targets, we performed proteomic profiling on serum samples collected at diagnosis using the O-Link Explore 384 Inflammation I panel. Among the 384 proteins analysed, 54 were differentially expressed between patients with and without cachexia (p < 0.05; **Figure 4A**, Supporting information **Table S2**), with 34 proteins upregulated and 20 downregulated in the cachexia group. After adjusting for multiple comparisons using Benjamini-Hochberg procedure, 16 proteins remained significantly different, including CCL23, IL-6, MLN, C1QA, AGRP and CKAP4 which remained significantly upregulated.

**Figure 4.**
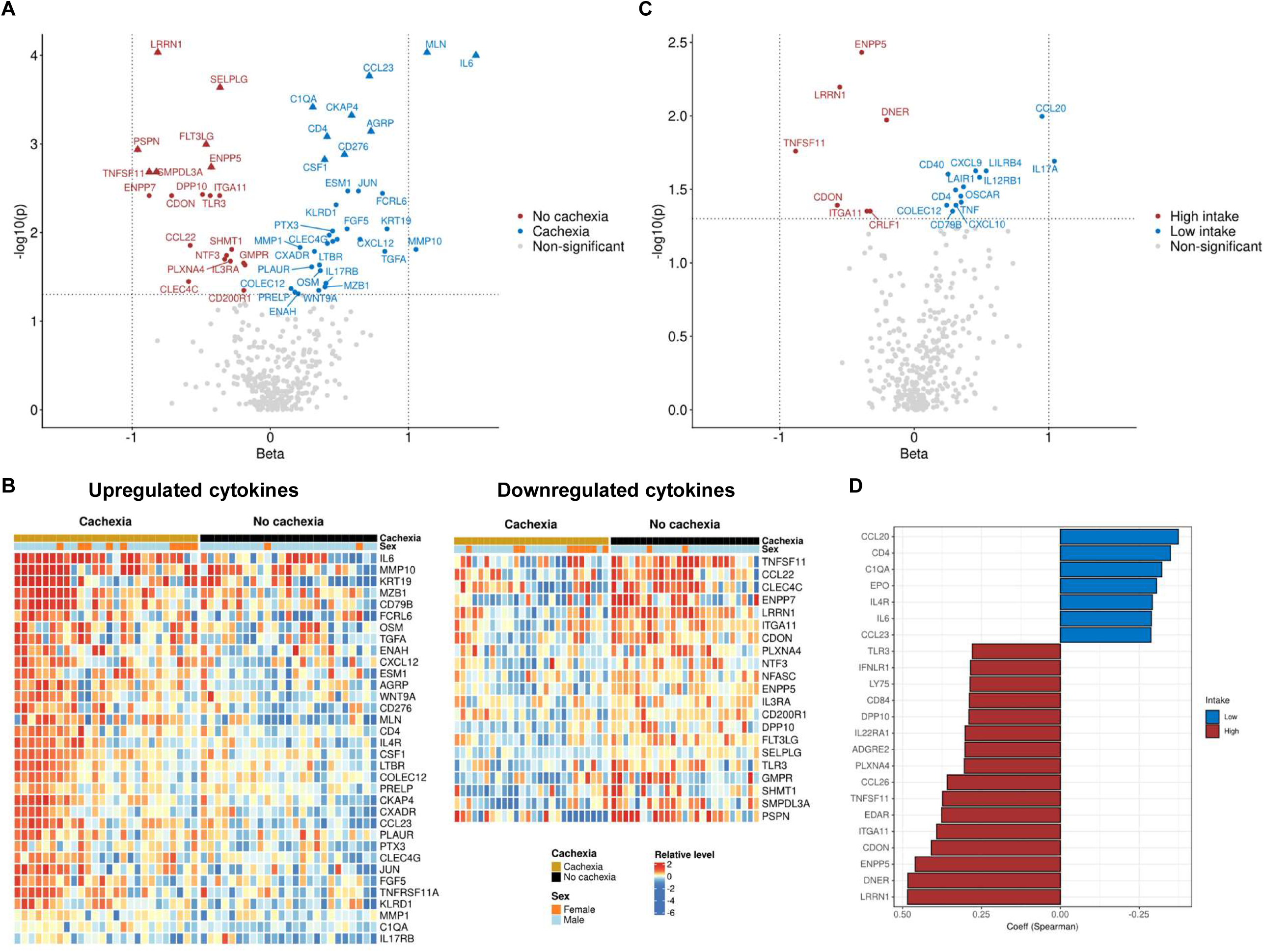
Serum proteome in patients with cachexia. (A) Volcano plot showing differentially expressed proteins between patients with cachexia (n = 27) and those without cachexia (n = 24). Proteins with nominal p-values < 0.05 are highlighted: red = associated with cachexia; blue = associated with no cachexia. Proteins significantly associated after p-value correction (FDR) are indicated with a triangle shape. (B) Unsupervised hierarchical clustering heatmaps of significantly upregulated (left) and downregulated (right) cytokines. Each column represents an individual patient, and each row corresponds to a protein. Colour scale indicates normalised protein abundance (NPX): red = higher, blue = lower. (C) Volcano plot showing differential protein abundance between patients with low (n=25) and high food intake (n=25) defined as those with reported intake above or below the median (1819.04 calories). Proteins with nominal p-values < 0.05 are highlighted: red = low intake; blue = high intake. (D) Bar plot showing proteins significantly associated with patient intake based on Spearman’s rank correlation.

Two members of the IL6 family, IL-6, and oncostatin M (OSM), as well as pentraxin-related protein 3 (PTX3), were among the 21 upregulated proteins based on nominal p-values. These proteins have previously been implicated as mediators of cancer cachexia in both preclinical models and clinical studies (17,18). Interestingly, other proteins previously associated with cancer cachexia, such as TNF-□, IL-1□, IL-8 (CXCL8), and VEGF, did not show significant differences between patients with or without cachexia in our cohort (**Table S2**).

Analysis of cachexia-associated cytokines revealed distinct co-secretion patterns (**Figure 4B**). IL-6 was elevated in most cachectic patients but were also increased in a subset of non-cachectic individuals. This finding highlights substantial interpatient variability in circulating markers and underscores the need for more precise patient stratification when developing targeted therapeutic interventions for cancer cachexia.

We also examined the correlation between significantly altered proteins and various cachexia-associated parameters, including percentage of weight loss, body composition, and caloric intake (**Fig. 4C and 4D**, **Tables 4 and 5**). Among the proteins upregulated in patients with cachexia, IL-6 and C-C motif chemokine ligand 23 (CCL23) were positively correlated with the degree of weight loss (**Table 4**). Notably, CCL23 and IL-6 also showed a moderate negative correlation with lower caloric intake (Spearman coefficient = −0.4, unadjusted p = 0.03 and coefficient = −0.29, unadjusted p=0.042, respectively).

**Table 4.**
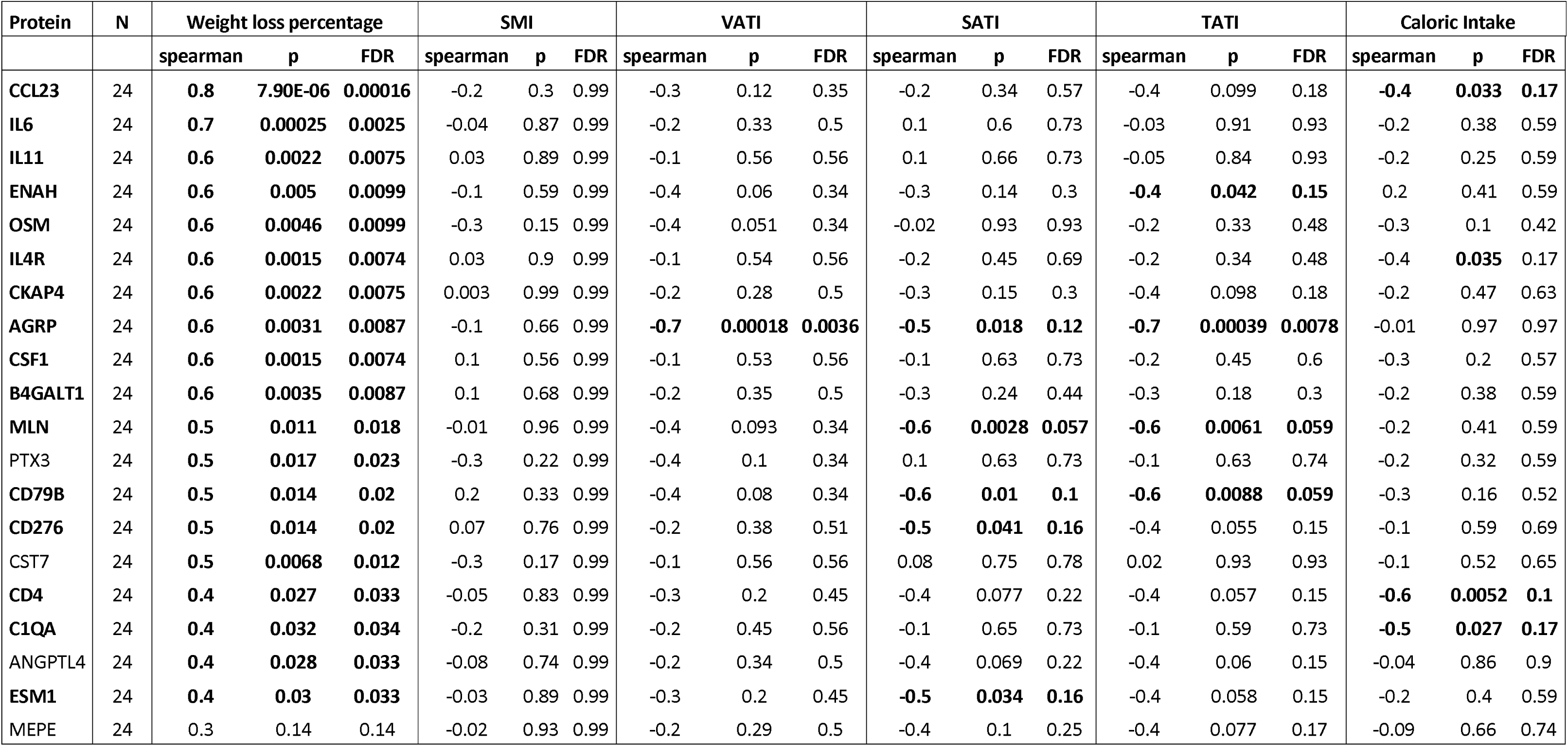
Correlation of upregulated proteins in patients with cachexia vs no cachexia with weight loss percentage and other cachexia-related parameters.

**Table 5.** Correlation of proteins downregulated in patients with cachexia with weight loss percentage and other cachexia-related parameters.

| Protein | N | Weight loss percentage |  |  | Skeletal muscle |  |  | Visceral adipose tissue |  |  | Subcutaneous adipose tissue |  |  | Total adipose tissue |  |  | Caloric Intake |  |  |
| --- | --- | --- | --- | --- | --- | --- | --- | --- | --- | --- | --- | --- | --- | --- | --- | --- | --- | --- | --- |
|  |  | spearman | p | FDR | spearman | p | FDR | spearman | p | FDR | spearman | p | FDR | spearman | p | FDR | spearman | p | FDR |
| DPP10 | 24 | **-0.7** | **0.00028** | **0.0041** | 0.3 | 0.14 | 0.51 | 0.1 | 0.61 | 0.95 | 0.2 | 0.36 | 0.86 | 0.3 | 0.26 | 0.92 | 0.4 | 0.051 | 0.28 |
| TNFSF11 | 24 | **-0.7** | **0.00058** | **0.0056** | -0.3 | 0.13 | 0.51 | 0.05 | 0.81 | 0.96 | 0.2 | 0.47 | 0.86 | 0.1 | 0.57 | 0.92 | 0.3 | 0.16 | 0.28 |
| CDON | 24 | **-0.7** | **0.00022** | **0.0041** | 0.1 | 0.59 | 0.85 | 0.3 | 0.25 | 0.95 | -0.1 | 0.52 | 0.86 | 0.02 | 0.93 | 0.96 | 0.3 | 0.1 | 0.28 |
| LRRN1 | 24 | **-0.6** | **0.0014** | **0.0091** | 0.04 | 0.88 | 0.94 | 0.06 | 0.79 | 0.96 | -0.1 | 0.62 | 0.86 | 0.01 | 0.96 | 0.96 | **0.4** | **0.029** | **0.28** |
| NTF3 | 24 | **-0.6** | **0.0025** | **0.01** | 0.3 | 0.15 | 0.51 | 0.02 | 0.93 | 0.96 | -0.3 | 0.15 | 0.86 | -0.1 | 0.56 | 0.92 | 0.4 | 0.065 | 0.28 |
| PTPRM | 24 | **-0.6** | **0.0035** | **0.013** | 0.2 | 0.3 | 0.72 | 0.2 | 0.49 | 0.95 | -0.009 | 0.97 | 0.97 | 0.1 | 0.67 | 0.92 | 0.2 | 0.36 | 0.44 |
| SMPDL3A | 24 | **-0.6** | **0.0019** | **0.0091** | -0.03 | 0.91 | 0.94 | 0.4 | 0.093 | 0.95 | 0.4 | 0.093 | 0.86 | 0.4 | 0.062 | 0.7 | 0.3 | 0.22 | 0.32 |
| ENPP5 | 24 | **-0.6** | **0.0018** | **0.0091** | **0.6** | **0.004** | **0.12** | 0.4 | 0.082 | 0.95 | 0.1 | 0.62 | 0.86 | 0.3 | 0.25 | 0.92 | 0.3 | 0.16 | 0.28 |
| IL1RL2 | 24 | **-0.5** | **0.024** | **0.05** | 0.06 | 0.79 | 0.93 | 0.3 | 0.14 | 0.95 | 0.3 | 0.2 | 0.86 | 0.4 | 0.071 | 0.7 | 0.2 | 0.4 | 0.46 |
| CNTNAP2 | 24 | **-0.5** | **0.014** | **0.035** | 0.1 | 0.54 | 0.85 | -0.1 | 0.65 | 0.95 | -0.1 | 0.59 | 0.86 | -0.05 | 0.84 | 0.96 | 0.04 | 0.86 | 0.89 |
| TNFSF10 | 24 | **-0.5** | **0.0059** | **0.019** | -0.4 | 0.11 | 0.51 | 0.02 | 0.92 | 0.96 | 0.2 | 0.36 | 0.86 | 0.1 | 0.53 | 0.92 | 0.3 | 0.18 | 0.28 |
| TPSAB1 | 24 | **-0.5** | **0.011** | **0.03** | 0.3 | 0.23 | 0.66 | 0.1 | 0.58 | 0.95 | 0.2 | 0.38 | 0.86 | 0.2 | 0.38 | 0.92 | 0.3 | 0.13 | 0.28 |
| FLT3LG | 24 | **-0.5** | **0.022** | **0.049** | 0.1 | 0.62 | 0.86 | 0.2 | 0.31 | 0.95 | 0.1 | 0.53 | 0.86 | 0.2 | 0.35 | 0.92 | 0.3 | 0.17 | 0.28 |
| PRSS8 | 24 | **-0.5** | **0.0066** | **0.019** | 0.3 | 0.14 | 0.51 | 0.04 | 0.88 | 0.96 | -0.2 | 0.3 | 0.86 | -0.1 | 0.67 | 0.92 | 0.3 | 0.1 | 0.28 |
| IL22RA1 | 24 | -0.4 | 0.088 | 0.11 | 0.06 | 0.78 | 0.93 | -0.1 | 0.63 | 0.95 | 0.009 | 0.97 | 0.97 | -0.03 | 0.9 | 0.96 | **0.5** | **0.01** | **0.28** |
| IL3RA | 24 | -0.4 | 0.063 | 0.089 | -0.06 | 0.79 | 0.93 | -0.2 | 0.36 | 0.95 | -0.2 | 0.46 | 0.86 | -0.2 | 0.36 | 0.92 | -0.003 | 0.99 | 0.99 |
| IL1A | 24 | -0.4 | 0.086 | 0.11 | 0.2 | 0.49 | 0.83 | 0.2 | 0.44 | 0.95 | -0.09 | 0.71 | 0.93 | 0.08 | 0.73 | 0.92 | 0.4 | 0.071 | 0.28 |
| PSPN | 24 | **-0.4** | **0.047** | **0.075** | **0.4** | **0.044** | **0.51** | 0.3 | 0.17 | 0.95 | 0.009 | 0.97 | 0.97 | 0.2 | 0.42 | 0.92 | 0.2 | 0.46 | 0.51 |
| SELPLG | 24 | -0.4 | 0.085 | 0.11 | 0.4 | 0.1 | 0.51 | 0.3 | 0.15 | 0.95 | 0.2 | 0.34 | 0.86 | 0.3 | 0.16 | 0.91 | 0.3 | 0.13 | 0.28 |
| CD83 | 24 | **-0.4** | **0.029** | **0.053** | 0.06 | 0.81 | 0.93 | 0.04 | 0.88 | 0.96 | -0.02 | 0.93 | 0.97 | 0.03 | 0.91 | 0.96 | 0.1 | 0.61 | 0.65 |
| ITGA11 | 24 | **-0.4** | **0.036** | **0.062** | 0.2 | 0.36 | 0.74 | 0.06 | 0.79 | 0.96 | -0.3 | 0.22 | 0.86 | -0.1 | 0.63 | 0.92 | 0.4 | 0.088 | 0.28 |
| RAB6A | 24 | -0.4 | 0.065 | 0.089 | 0.001 | 1 | 1 | 0.2 | 0.39 | 0.95 | 0.4 | 0.06 | 0.86 | 0.4 | 0.097 | 0.7 | 0.2 | 0.3 | 0.42 |
| **SHMT1** | 24 | **-0.4** | **0.029** | **0.053** | -0.2 | 0.41 | 0.79 | 0.1 | 0.59 | 0.95 | 0.3 | 0.17 | 0.86 | 0.3 | 0.22 | 0.92 | 0.3 | 0.11 | 0.28 |
| **CCL17** | 24 | -0.4 | 0.091 | 0.11 | 0.04 | 0.85 | 0.94 | -0.1 | 0.52 | 0.95 | -0.02 | 0.95 | 0.97 | -0.1 | 0.64 | 0.92 | **0.4** | **0.044** | **0.28** |
| **CCL22** | 24 | **-0.4** | **0.049** | **0.075** | -0.2 | 0.34 | 0.74 | -0.2 | 0.51 | 0.95 | -0.04 | 0.86 | 0.97 | -0.09 | 0.68 | 0.92 | 0.2 | 0.33 | 0.43 |
| ALDH3A1 | 24 | -0.3 | 0.12 | 0.13 | 0.3 | 0.16 | 0.51 | -0.02 | 0.93 | 0.96 | -0.2 | 0.47 | 0.86 | -0.1 | 0.64 | 0.92 | 0.2 | 0.35 | 0.44 |
| ITGA6 | 24 | -0.3 | 0.1 | 0.11 | 0.3 | 0.27 | 0.7 | 0.3 | 0.24 | 0.95 | 0.3 | 0.15 | 0.86 | 0.4 | 0.097 | 0.7 | 0.3 | 0.18 | 0.28 |
| **PLXNA4** | 24 | -0.3 | 0.15 | 0.15 | 0.2 | 0.47 | 0.83 | 0.1 | 0.6 | 0.95 | -0.08 | 0.74 | 0.93 | 0.04 | 0.85 | 0.96 | **0.4** | **0.035** | **0.28** |
| DECR1 | 24 | -0.2 | 0.4 | 0.4 | 0.1 | 0.56 | 0.85 | 0.001 | 1 | 1 | 0.2 | 0.47 | 0.86 | 0.08 | 0.73 | 0.92 | 0.3 | 0.16 | 0.28 |

The neuropeptide agouti-related protein (AGRP), known to stimulate food intake, was paradoxically elevated in cachectic patients, and it was inversely correlated with VATI (Spearman coefficient = −0.7, p = 0.0036) and TATI (Spearman coefficient = −0.7, p = 0.0078). Other proteins like CD4 and the IL-4 receptor, although not secreted proteins, were detected in the blood of patients with cachexia and with lower food intake (**Table 4**).

We examined cytokines that were reduced in the blood of cachectic patients compared to non-cachectic patients and identified several proteins whose levels inversely correlated with the percentage of weight loss or with food intake (**Table 5**, **Fig 4C and 4D**). Among these, TNFSF11 (RANK ligand) was lower in cachectic patients and significantly decreased as weight loss became more pronounced and the estimated food intake was lower, while its receptor RANK (TNFRSF11) was positively associated with cachexia. LRRN1, involved in neuronal development and cell–cell interactions, was also correlated with food intake. Other proteins associated with intake were ENPP5, CDON, ITGA11, PLXNA4, DPP10, and TLR3 (**Table 5**). Analysis of associations with food intake independently of cachexia also showed association of higher CCL20 and IL17A with lower intake (**Fig. 4D**).

## DISCUSSION

Although the field of cancer cachexia is advancing rapidly, important questions remain about the underlying mechanisms and factors driving this complex syndrome. In lung cancer, most previous studies have focused on the association of cachexia with body composition or selected circulating cytokines. To date, few studies have focused on locally advanced NSCLC and have adopted a multidisciplinary approach that simultaneously integrates nutritional status, circulating biomarkers, and body composition within a prospective study design. In addition, most prior research has primarily examined the evolution of cachexia during treatment and its impact on treatment outcomes. The aim of the present study was to determine the prevalence of lung cancer-associated cachexia in a prospective cohort of patients with locally advanced NSCLC prior to initiation of the oncological treatment. In addition, we sought to comprehensively characterize the pre-treatment cachexia phenotype across multiple dimensions, including its association with body composition, clinicopathological features, metabolic parameters from blood tests, nutritional status, and physical function, and circulating cytokines assessed through proteomic analysis.

In our study, more than half (53%) of the patients met the criteria for cachexia, according to Fearon’s definition. This high prevalence is consistent with previous reports in both early and advanced NSCLC (19). Cachexia has been associated with unfavorable survival outcomes, underscoring the importance of early identification, particularly in the curative setting of lung cancer. Early diagnosis may enable timely interventions that could improve clinical management and patient outcomes.

Although most female participants in our study were classified within the cachexia group, this finding should be interpreted with caution due to the underrepresentation of women in the cohort. This imbalance reflects the higher prevalence of locally advanced NSCLC among males in our country. Validation of this observation in larger, more gender-balanced cohorts is warranted and is currently ongoing. An important observation was that 41% of patients classified as cachectic were either overweight or obese, underscoring the limitation of relying solely on BMI for nutritional analysis. Our multidimensional profiling further revealed that, although cachexia was associated with lower skeletal muscle index (SMI), a substantial proportion of cachectic patients did not exhibit low muscle mass at baseline. Likewise, despite overall correlations with lower adipose tissue and BMI, several patients meeting cachexia criteria maintained preserved fat and muscle compartments. These findings highlight a disconnect between conventional cachexia definitions, primarily based on weight loss, and the underlying alterations in body composition. This observation underscores the limitations of static, cross-sectional assessments and supports the need for longitudinal, individualized monitoring to better capture the dynamic nature of cachexia and inform personalized, multimodal interventions.

In addition to involuntary weight loss and depletion of muscle and adipose tissue, cancer cachexia is characterized at the molecular level by host metabolic dysregulation and systemic inflammation. In our study, cachectic patients exhibited elevated levels of CRP, a well-established marker of inflammation. This result aligns with previous reports and further supports the link between cachexia and inflammatory processes (20,21). From a nutritional standpoint, cachexia was strongly associated with moderate to severe malnutrition, whereas nearly all patients classified as non-cachectic displayed adequate nutritional status. Notably, cachectic patients reported significantly lower fat intake, based on a 24h dietary recall, while no statistical differences were observed for protein, carbohydrate or fiber consumption. We speculate that the higher prevalence of anorexia among cachectic patients may contribute to a reduced preference for high-fat foods. Whether this altered intake is modulated by specific hormones or tumour-related cytokines acting at the level of the central nervous system remains unknown. Reduced fat intake could potentially accelerate adipose tissue depletion; however, further studies are needed to confirm and better elucidate these observations.

Several recent studies in animal models have demonstrated the role of tumour-derived factors, such as lactate or cytokines, in promoting tumor progression, disrupting host metabolism, and contributing to the development of cancer cachexia (17,22,23). We assessed the association between cachexia and several previously described cytokines, including GDF-15 and members of the IL-6 family in our cohort prior to initiation of oncologic treatment. Our findings confirmed that some of these cytokines were significantly elevated in patients classified in the cachexia group, consistent with prior observations in other cancer populations. However, other cytokines traditionally linked to cachexia, such as TNF-□or IL-8, were not elevated in our cohort. These findings suggest that the contribution of specific cytokines may be tumor-type or stage-dependent. Moreover, we observed substantial interpatient variability in circulating cytokines among patients with cachexia, underscoring the need for precision medicine approaches in the study and management of this syndrome, including biomarker-guided patient selection for targeted interventions.

We observed that GDF-15 elevation was common among patients with locally advanced NSCLC and was significantly associated with the percentage of weight loss. However, its low specificity limits its utility as a standalone marker for diagnosing pre-treatment cachexia in this population. IL-6, consistently associated with cachexia in both clinical and preclinical studies (24–26) was also elevated. Certain cytokines exhibited an intriguing expression pattern, including agouti-related protein (AGRP). Although AGRP promotes appetite and has been shown to attenuate cachectic phenotypes in preclinical models of colon cancer (27), it was paradoxically elevated in patients with cachexia in our cohort. The inverse relationship between AGRP levels and adipose tissue index suggests that its upregulation may represent a compensatory response to weight loss and fat depletion. Importantly, the simultaneous elevation of both pro-cachectic and anti-cachectic cytokines highlights the complex interplay between inflammatory mediators, neuropeptides, and alterations body composition that occur during progression of cancer cachexia. Another protein showing an intriguing association with cachexia was the digestion hormone Motilin (MLN), which has not been directly linked to cachexia but is associated with hunger, and agonists of its receptor have been reported to increase appetite (28).

CCL23 has been associated with post-surgical muscle loss in humans but has not previously been linked with cachexia (29). C1QA is a component of the C1 complex of the complement system, and cachexia is strongly associated with systemic inflammation. Therefore, the association between C1QA and cachexia may reflect activation of the complement cascade, consistent with the inflammatory state characteristic of this syndrome (30). CCL23, motilin, and IL-6 were the cytokines that showed the strongest correlation with cachexia, and the first two have not previously been associated with this syndrome, highlighting their potential relevance as novel targets. The remaining proteins that remained significant after correction (CD276, CD4, CSF-1, and CKAP4) may also be relevant in the context of cachexia, possibly reflecting underlying inflammatory processes. Other cytokines previously implicated in cancer-associated cachexia across different tumor types were also differentially expressed in our cohort, including Pentraxin 3 (PTX3). PTX3 has been reported to be elevated in the serum of patients with pancreatic cancer experiencing weight loss (18) as well as in patients with NSCLC-associated cachexia treated with immunotherapy (20), supporting its potential as a candidate for further investigation. Another IL-6 superfamily member was also upregulated, Oncostatin M (OSM), which has demonstrated muscle-wasting effects *in vivo* (31,32). Notably, our study also identified elevated levels of several cytokines not previously associated with cachexia: endothelial cell-specific molecule 1 (ESM1), and enabled homolog (ENAH), none of which, to our knowledge, have been previously implicated in cancer cachexia. These newly identified cytokines represent promising candidates for further investigation as potential biomarkers of cancer cachexia. The causal association between these upregulated proteins with cachexia remains to be determined in experimental models. Conversely, some cytokines previously reported to be positively associated with cachexia, such as RANK ligand, which plays a role in inflammation and bone resorption, particularly in women and female mouse models (33), were found to be decreased in our predominantly male cachectic population.

Our study has some limitations primarily related to the limited sample size, which reduces the statistical power of our analyses. However, the prospective and longitudinal design, along with the comprehensive multidisciplinary assessment, adds significant value to our findings. We plan to validate these results in a larger cohort and address the underrepresentation of women observed in the present study. Current diagnostic criteria for cancer cachexia, as proposed by Fearon et al. in 2011, primarily rely on weight loss, and may not fully capture the complexity of the syndrome in light of recent advances in the field. While definitions for sarcopenia and adipose tissue depletion have been proposed, universally accepted or standardised cut-off values are still lacking, especially in oncologic populations. In our study, low SMI was defined using sex-specific tertiles, but this approach requires further validation. Additionally, body composition data were derived from routine CT scans, which may not be consistently available for all patients prior to treatment, potentially limiting accurate quantification of sarcopenia.

In conclusion, this prospective study demonstrates that cachexia is highly prevalent at diagnosis in patients with unresectable locally advanced NSCLC. To the best of our knowledge, no previous studies in this clinical setting have integrated all the dimensions assessed here, including body composition, weight loss, nutritional assessment, physical function, clinical covariates, and proteomics profiling within the same population. Despite significant challenges, we believe this work is clinically relevant and contributes to advancing the field towards improved patient stratification and more tailored treatment strategies.

## Supporting information

Hijazo-Pechero supporting information - Table S1

Hijazo-Pechero supporting information - Table S2

## Data Availability

All data produced in the present study are available upon reasonable request to the authors.

## Acknowledgements

We thank Dolors Dot Bach from the Laboratori Clínic Territorial Metropolitana Sud from the Hospital Universitari de Bellvitge, for help with GDF-15 analysis. Isabel Brao and Monica Arellano for help attending patients.

## Ethical standards

Human studies have been approved by the Bellvitge ethics committee and have therefore been performed in accordance with the ethical standards laid down in the 1964 Declaration of Helsinki and its later amendments.

---

**Sup. Fig. 1.**
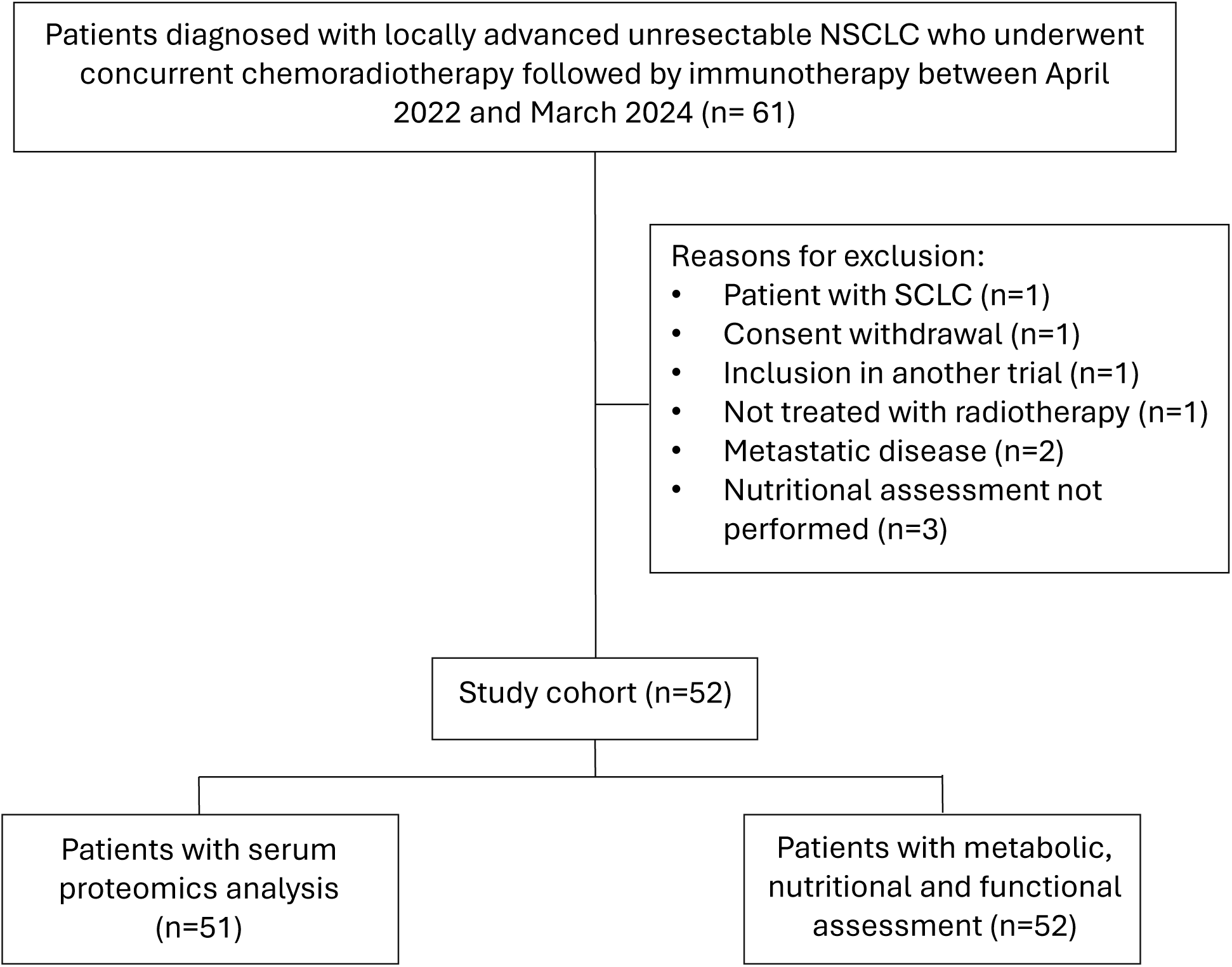
Flow diagram

